# COVID-19 in breast cancer patients: a cohort at the Institut Curie hospitals in the Paris area

**DOI:** 10.1101/2020.04.30.20085928

**Authors:** Perrine Vuagnat, Maxime Frelaut, Toulsie Ramtohul, Clémence Basse, Sarah Diakite, Aurélien Noret, Audrey Bellesoeur, Vincent Servois, Delphine Hequet, Enora Laas, Youlia Kirova, Luc Cabel, Jean-Yves Pierga, Institut Curie Breast Cancer and COVID Group, Laurence Bozec, Xavier Paoletti, Paul Cottu, François-Clément Bidard

**Author notes:** equal contribution (Laurence Bozec, Xavier Paoletti, Paul Cottu, François-Clément Bidard).

## Abstract

**Background:** Cancer patients have been reported to be at higher risk of COVID-19 complications and deaths. We report the characteristics and outcome of patients diagnosed with COVID-19 during breast cancer treatment at Institut Curie hospitals (ICH, Paris area, France).

**Methods:** An IRB-approved prospective registry was set up at ICH on March 13^th^, 2020 for all breast cancer patients with COVID-19 symptoms or radiologic signs. Registered data included patient history, tumor characteristics and treatments, COVID-19 symptoms, radiological features and outcome. Data extraction was done on April 25^th^, 2020. COVID-19 patients were defined as those with either a positive RNA test or typical, newly appeared lung CT-scan abnormalities.

**Results:** Among 15,600 patients actively treated for early or metastatic breast cancer during the last 4 months at ICH, 76 patients with suspected COVID-19 infection were included in the registry and followed. Fifty-nine of these patients were diagnosed with COVID-19 based on viral RNA testing (N=41) or typical radiologic signs: 37/59 (63%) COVID-19 patients were treated for metastatic breast cancer, and 13/59 (22%) of them were taking corticosteroids daily. Common clinical features mostly consisted of fever and/or cough, while ground-glass opacities were the most common radiologic sign at diagnosis. We found no association between prior radiation therapy fields or extent of radiation therapy sequelae and extent of COVID-19 lung lesions. Twenty-eight of these 59 patients (47%) were hospitalized and 6 (10%) were transferred to an intensive care unit. At the time of analysis, 45/59 (76%) patients were recovering or had been cured, 10/59 (17%) were still followed and 4/59 (7%) had died from COVID-19. All 4 patients who died had significant non-cancer comorbidities. In univariate analysis, hypertension and age (>70) were the two factors associated with a higher risk of intensive care unit admission and/or death.

**Conclusions:** This prospective registry analysis suggests that the COVID-19 mortality rate in breast cancer patients depends more on comorbidities than prior radiation therapy or current anti-cancer treatment. Special attention must be paid to comorbidities when estimating the risk of severe COVID-19 in breast cancer patients.

## Background

On December 31^st^, 2019, the World Health Organization was informed about cases of pneumonia of unknown cause in Wuhan, China [1]. A novel coronavirus, named severe acute respiratory syndrome coronavirus 2 (SARS-CoV-2), was identified as the cause of coronavirus disease 2019 (COVID-19) [2]. Over the following months, the viral outbreak shifted from China to the rest of the world and was subsequently recognized as a pandemic [3]. In France, the number of confirmed cases rose during early March: more than 2,800 confirmed cases were officially reported on March 13, 2020, when the French hospital emergency response plan, which coordinates all hospitals, was increased to its maximum readiness level [4]. As of April 24, France is the 6^th^ most severely affected country in the world (in terms of absolute numbers), with more than 21,000 official COVID-19-related deaths [4, 5]. With about 6,000 deaths, the Paris area is one of the most severely affected regions in France [6].

The first report on COVID-19 outcome in cancer patients was published on February 14^th^ [7]: in a series of 18 Chinese patients with a history of cancer and a diagnosis of COVID-19, 7 (39%) had to be treated in the intensive care unit (ICU) and/or died. This seminal retrospective study prompted major concerns about the risk of COVID-19 infection in cancer patients. Further studies confirmed that, compared to the Chinese general population, cancer patients are at higher risk of severe COVID-19 symptoms and death [7-10]. Cancer patients with blood, lung or metastatic cancers were reported to have the highest frequency of severe outcome [8, 10]. These retrospective reports, of limited size and restricted to patients hospitalized in Chinese hospitals, may not be fully transposable to Western healthcare systems, as suggested by a preliminary report on New Yorkers admitted to ICU [11].

Our study reports the COVID-19 features and outcomes experienced by inpatients and outpatients actively treated for breast cancer at Institut Curie hospitals (ICH) in the Paris area, France.

## Methods

### Registry

The prospective COVID-19 registry was approved by the ICH institutional review board, which waived documentation of informed consent due to its observational nature. Starting from March 13th, 2020, all proven or suspected COVID-19 cases were prospectively registered. Declaration of all proven or suspected cases was made mandatory by the ICH Director, and done by any doctor or nurse to a unique email address, even if RNA test was not done or available. Emails were checked several times a day by a team of 5 doctors; all declared patients were included in the registry on the same day (day 1) and followed-up. The standardized follow-up included phone calls to patients which were scheduled at days 8, 14 and 28 and tracked in the central registry. Follow-up calls were initially given by doctors, later joined by other ICH qualified healthcare workers (such as genetic counsellors, who received a training on COVID-19) for patients that had mild symptoms or who were recovering. More frequent and/or longer follow-up was provided whenever medically necessary. Patients hospitalized outside IC hospitals were also registered and prospectively followed. The list of patients who had an RNA test prescribed at ICH was also investigated (with no missing case identified). Data captured in the registry are displayed in **Supplementary Methods 1**. For this analysis, data were extracted on April 25^th^, 2020. This report was written according to the STROBE checklist.

### Breast cancer care at IC during the SARS-CoV-2 pandemic

Guidances on breast cancer care during the pandemic are detailed in **Supplementary Methods 2**. *COVID-19 diagnosis: laboratory tests and radiology*

#### SARS-CoV-2 RNA tests

Testing was initially restricted to critically ill patients with COVID-19 symptoms, but subsequently became available to all cancer patients (including outpatients that were under active treatment) with suspected COVID-19 at the end of March 2020. Nasopharyngeal swabs were analyzed for SARS-CoV-2 RNA by reverse-transcription polymerase chain reaction assays targeting 2 regions of the viral RdRp gene. All assays used in France had to be validated by the French National Reference Center (Institut Pasteur, Paris, France) [12, 13].

#### CT scan protocol and image interpretation

Whenever available, CT images were centrally reviewed by two senior radiologists with consensus qualitative and semiquantitative assessment. In accordance with previous reports on COVID-19 imaging [14, 15], the following patterns were sought: ground-glass opacity, crazy paving (ground glass opacity associated with interlobular septal thickening [16]), focal consolidation and linear consolidation. To be included in the COVID-19 population, patients with negative or not available RNA test had to display typical and newly acquired (i.e. not pre-existing on the previous CT-scan) COVID-19 related lung lesions. The predominant pattern was determined for each examination. The severity (%) of lung involvement was evaluated according to the French Society of Radiology guidelines [17]. The presence of lung or pleural metastases was assessed by comparison with previous CT scans. Lung radiation therapy sequelae were evaluated by semiquantitative evaluation of confluent radiologic opacities (grade 3 of the Lent-Soma scoring system [18]) affecting the right, left or middle lobes (no involvement; ≤10% of lung volumes; 11-25%; ≥26%).

### Statistics

The main study population, “COVID-19 patients”, is defined as those with positive RNA test or for whom RT-PCR result was not available (or pending) but who had suggestive radiologic findings. We also report data on the subgroup of patients who had biological confirmation of COVID-19 status using RT-PCR, referred to as “RNA-positive subgroup”, but did not perform statistical analyses on that subgroup. Main outcome of patients was defined as death or ICU admission. Descriptive and univariate prognostic factor analysis was performed. Two sensitivity analyses were performed: (i) using death only and (ii) using time to death or ICU admission to account for patients with partial follow-up. As sensitivity analyses, prognostic factor analysis of death only. Due to the highly explorative nature of the report and the small number of events, no adjustment for multiple testing was applied and multivariate analysis was not done. All analyses were performed in SAS v9.4 and R software.

## Results

### COVID-19 diagnosis

From March 13^th^, 2020 to April 25^th^, 2020 (date of data extraction), 76 patients actively treated for breast cancer were included in the ICH COVID-19 registry. For comparison, 15,600 breast cancer patients had at least one consultation or treatment for breast cancer at one of the IC hospitals in the 4 months before lockdown (11/01/2019 to 02/28/2020). The patient flow-chart for the COVID-19 registry is displayed in **Figure 1A**. RNA testing was performed in 58 patients, while CT scan was performed in 39 patients. A total of 59 patients were diagnosed with COVID-19, based on either a positive SARS-CoV-2 RNA test (N=41 patients; “RNA-positive subgroup”) or, in the case of negative or missing RNA test, radiologic findings (N=18 patients). Seventeen patients only reported symptoms suggestive of COVID-19 that were not confirmed by RNA test and/or lung CT scan. Most patients in the subgroup who underwent both RNA testing and CT scan presented concordant results, as displayed in **Figure 1B**.

**Figure 1:**
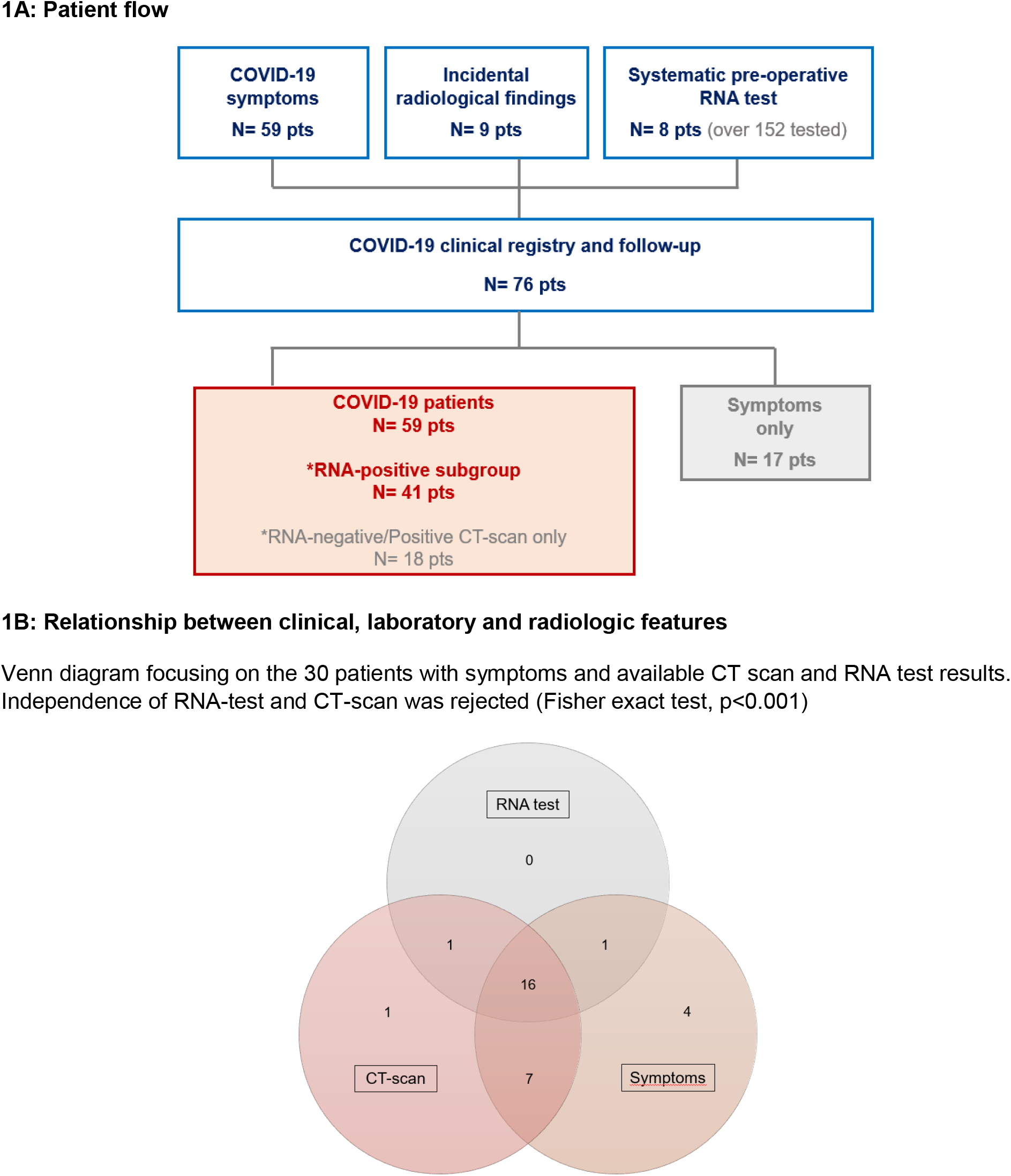
Patient flow and COVID-19 testing.

### Patient history

Breast cancer patient history and comorbidities are shown in **Table 1**. Ten of the 59 COVID-19 patients (17%) were older than 70. Other notable comorbidities among COVID-19 patients included hypertension (36%), obesity (17%), diabetes (17%) and heart disease (14%). The most frequent comedication in this population was corticosteroids (22%, defined as a daily intake of more than 20 mg-equivalent dose of prednisolone, excluding chemotherapy comedications). Reasons for corticosteroid intake were symptomatic brain or leptomeningeal metastasis (10%), epiduritis (5%), other cancer-related symptoms (5%) and autoimmune hepatitis (2%), respectfully. All these patients have been under corticosteroids for at least one month. About two-thirds of COVID-19 patients (and all those treated with corticosteroids) were treated for metastatic breast cancer. As shown in **Table 2**, ongoing anti-cancer treatments were representative of those currently administered to patients treated for early or metastatic disease, most commonly chemotherapy (49%), followed by endocrine therapy (32%).

**Table 1:**
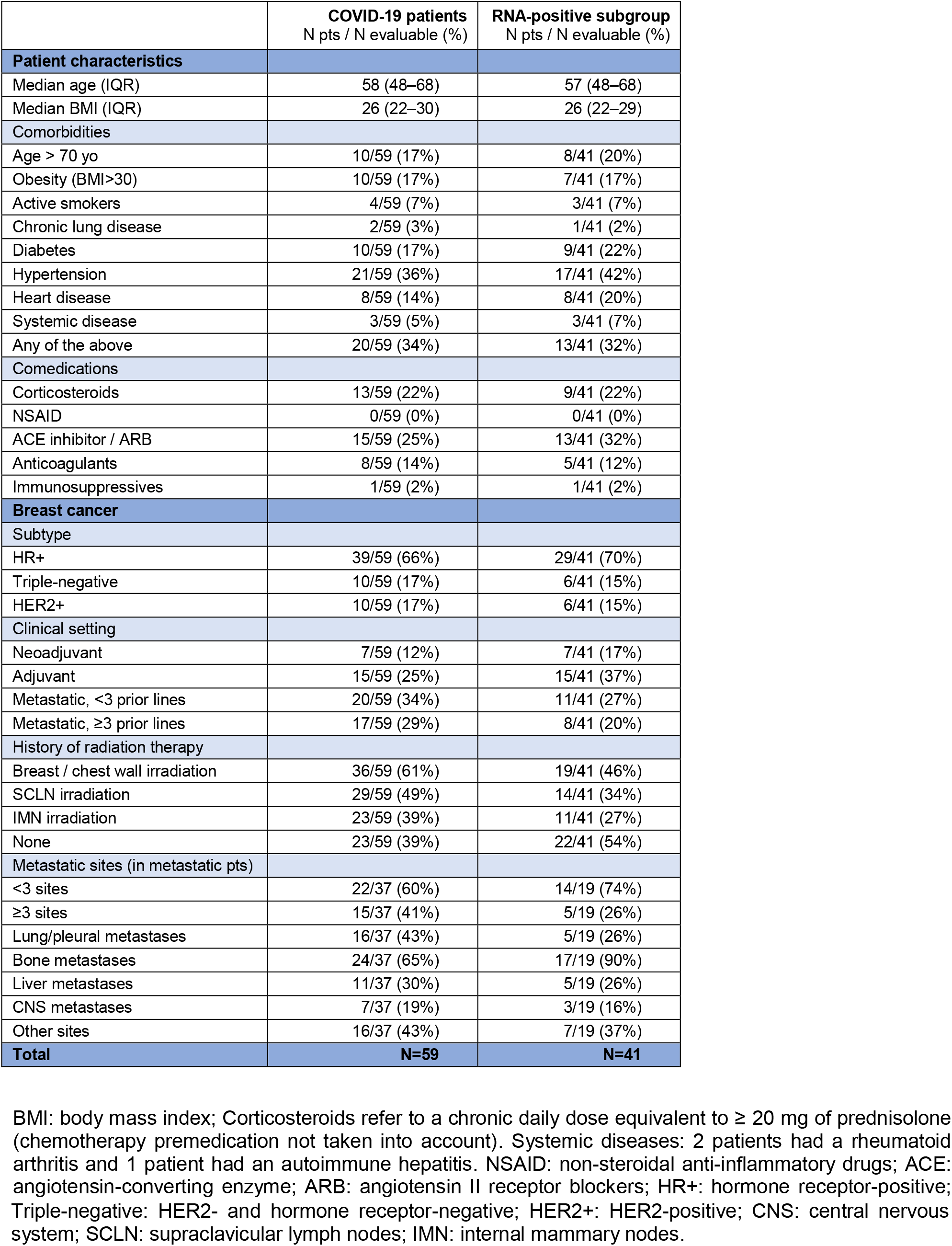
Patients’ medical history.

|  | **COVID-19 patients**<br>N pts / N evaluable (%) | **RNA-positive subgroup**<br>N pts / N evaluable (%) |
| --- | --- | --- |
| **Patient characteristics** |  |  |
| Median age (IQR) | 58 (48–68) | 57 (48–68) |
| Median BMI (IQR) | 26 (22–30) | 26 (22–29) |
| **Comorbidities** |  |  |
| Age > 70 yo | 10/59 (17%) | 8/41 (20%) |
| Obesity (BMI>30) | 10/59 (17%) | 7/41 (17%) |
| Active smokers | 4/59 (7%) | 3/41 (7%) |
| Chronic lung disease | 2/59 (3%) | 1/41 (2%) |
| Diabetes | 10/59 (17%) | 9/41 (22%) |
| Hypertension | 21/59 (36%) | 17/41 (42%) |
| Heart disease | 8/59 (14%) | 8/41 (20%) |
| Systemic disease | 3/59 (5%) | 3/41 (7%) |
| Any of the above | 20/59 (34%) | 13/41 (32%) |
| **Comedications** |  |  |
| Corticosteroids | 13/59 (22%) | 9/41 (22%) |
| NSAID | 0/59 (0%) | 0/41 (0%) |
| ACE inhibitor / ARB | 15/59 (25%) | 13/41 (32%) |
| Anticoagulants | 8/59 (14%) | 5/41 (12%) |
| Immunosuppressives | 1/59 (2%) | 1/41 (2%) |
| **Breast cancer** |  |  |
| **Subtype** |  |  |
| HR+ | 39/59 (66%) | 29/41 (70%) |
| Triple-negative | 10/59 (17%) | 6/41 (15%) |
| HER2+ | 10/59 (17%) | 6/41 (15%) |
| **Clinical setting** |  |  |
| Neoadjuvant | 7/59 (12%) | 7/41 (17%) |
| Adjuvant | 15/59 (25%) | 15/41 (37%) |
| Metastatic, <3 prior lines | 20/59 (34%) | 11/41 (27%) |
| Metastatic, ≥3 prior lines | 17/59 (29%) | 8/41 (20%) |
| **History of radiation therapy** |  |  |
| Breast / chest wall irradiation | 36/59 (61%) | 19/41 (46%) |
| SCLN irradiation | 29/59 (49%) | 14/41 (34%) |
| IMN irradiation | 23/59 (39%) | 11/41 (27%) |
| None | 23/59 (39%) | 22/41 (54%) |
| **Metastatic sites (in metastatic pts)** |  |  |
| <3 sites | 22/37 (60%) | 14/19 (74%) |
| ≥3 sites | 15/37 (41%) | 5/19 (26%) |
| Lung/pleural metastases | 16/37 (43%) | 5/19 (26%) |
| Bone metastases | 24/37 (65%) | 17/19 (90%) |
| Liver metastases | 11/37 (30%) | 5/19 (26%) |
| CNS metastases | 7/37 (19%) | 3/19 (16%) |
| Other sites | 16/37 (43%) | 7/19 (37%) |
| **Total** | **N=59** | **N=41** |

BMI: body mass index; Corticosteroids refer to a chronic daily dose equivalent to $\geq 20$ mg of prednisolone (chemotherapy premedication not taken into account). Systemic diseases: 2 patients had a rheumatoid arthritis and 1 patient had an autoimmune hepatitis. NSAID: non-steroidal anti-inflammatory drugs; ACE: angiotensin-converting enzyme; ARB: angiotensin II receptor blockers; HR+: hormone receptor-positive;
Triple-negative: HER2- and hormone receptor-negative; HER2+: HER2-positive; CNS: central nervous system; SCLN: supraclavicular lymph nodes; IMN: internal mammary nodes.

**Table 2:** Ongoing treatments.

|  | COVID-19 patients<br>N pts / N evaluable (%) | RNA-positive subgroup<br>N pts / N evaluable (%) |
| --- | --- | --- |
| **Early breast cancer patients** | **N=22** | **N=22** |
| Surgery* | 3/22 (14%) | 3/22 (14%) |
| Chemotherapy | 8/22 (36%) | 8/22 (36%) |
| Epirubicin and cyclophosphamide | 5/22 (23%) | 5/22 (23%) |
| Paclitaxel/Docetaxel | 2/22 (9%) | 2/22 (9%) |
| Radiation therapy | 2/22 (9%) | 2/22 (9%) |
| Endocrine therapy | 4/22 (18%) | 4/22 (18%) |
| Anti-oestrogens | 3/22 (14%) | 3/22 (14%) |
| Aromatase Inhibitors | 1/22 (4%) | 1/22 (4%) |
| Targeted therapy | 3/22 (14%) | 3/22 (14%) |
| Trastuzumab | 2/22 (9%) | 2/22 (9%) |
| Pertuzumab | 1/22 (4%) | 1/22 (4%) |
| Trastuzumab emtansine | 1/22 (4%) | 1/22 (4%) |
| None | 6/22 (27%) | 6/22 (27%) |
| Pending surgery | 5/22 (23%) | 5/22 (23%) |
| Pending radiation therapy | 1/22 (4%) | 1/22 (4%) |
| Combination of any treatment | 3/22 (14%) | 3/22 (14%) |
| **Metastatic breast cancer patients** | **N=37** | **N=19** |
| Surgery* | 0 (0%) | 0 (0%) |
| Chemotherapy | 21/37 (57%) | 10/19 (53%) |
| Capecitabine | 7/37 (19%) | 4/19 (21%) |
| Paclitaxel/Docetaxel | 6/37 (16%) | 3/19 (16%) |
| Epirubicin and cyclophosphamide | 1/37 (3%) | 0 (0%) |
| Vinorelbine | 2/37 (5%) | 0 (0%) |
| Eribulin | 1/37 (3%) | 1/19 (5%) |
| Gemcitabine | 2/37 (5%) | 1/19 (5%) |
| Carboplatin | 3/37 (8%) | 1/19 (5%) |
| Intrathecal chemotherapy | 2/37 (5%) | 2/19 (10%) |
| Radiation therapy | 2/37 (5%) | 2/19 (10%) |
| Endocrine therapy | 15/37 (40%) | 7/19 (37%) |
| Anti-oestrogens | 3/37 (8%) | 1/19 (5%) |
| Aromatase Inhibitors | 11/37 (30%) | 6/19 (32%) |
| Selective Estrogen Receptor Degradar | 1/37 (3%) | 0 (0%) |
| Targeted therapy | 16/37 (43%) | 7/19 (37%) |
| CDK4/6 inhibitor | 9/37 (24%) | 5/19 (26%) |
| Trastuzumab | 5/37 (13%) | 2/19 (10%) |
| Pertuzumab | 4/37 (11%) | 2/19 (10%) |
| Everolimus** | 2/37 (5%) | 0 (0%) |
| Immunotherapy | 0 (0%) | 0 (0%) |
| Combination of any treatment | 21/37 (57%) | 8/19 (42%) |
| None | 1/37 (3%) | 1/19 (5%) |
| **Total** | **N=59** | **N=41** |

Listed treatments were those ongoing within 30 days before COVID-19 diagnosis. \* Ongoing surgery includes 30 days from surgery. \*\* as recommended, patients stopped everolimus at the beginning of the pandemic.

### Features at diagnosis

Clinical, laboratory and radiologic features at diagnosis are displayed in **Table 3**. Fever and cough were the most common symptoms, observed in 46% and 37% of COVID-19 patients, respectively. Nine of the 59 patients (18%) developed COVID-19 symptoms more than 2 days after being admitted to hospital (IC or elsewhere), corresponding to the interval used to define nosocomial infections. The mean absolute lymphocyte count was normal (1.5/mm^3^). Most patients had no or limited extent of COVID-19 lung disease, as 25/28 patients (89%) had less than 25% involvement of their lung volume. Twenty-eight CT-scan were available for central review: the most common radiologic feature was ground-glass opacities, observed in 14/28 (50%) of COVID-19 patients with CT scan at diagnosis. No significant association was observed between these characteristics or the presence of lung metastases and the extent of COVID-19 lung disease. **Supplementary Figure 1** displays, for each COVID-19 patient, the prior radiation therapy fields, radiation therapy sequelae and extent of COVID-19 lung disease. There was no association between prior radiation therapy and the extent of COVID-19 lesions (≤10% vs >10%, Fisher exact test p=0.69).

**Table 3:**
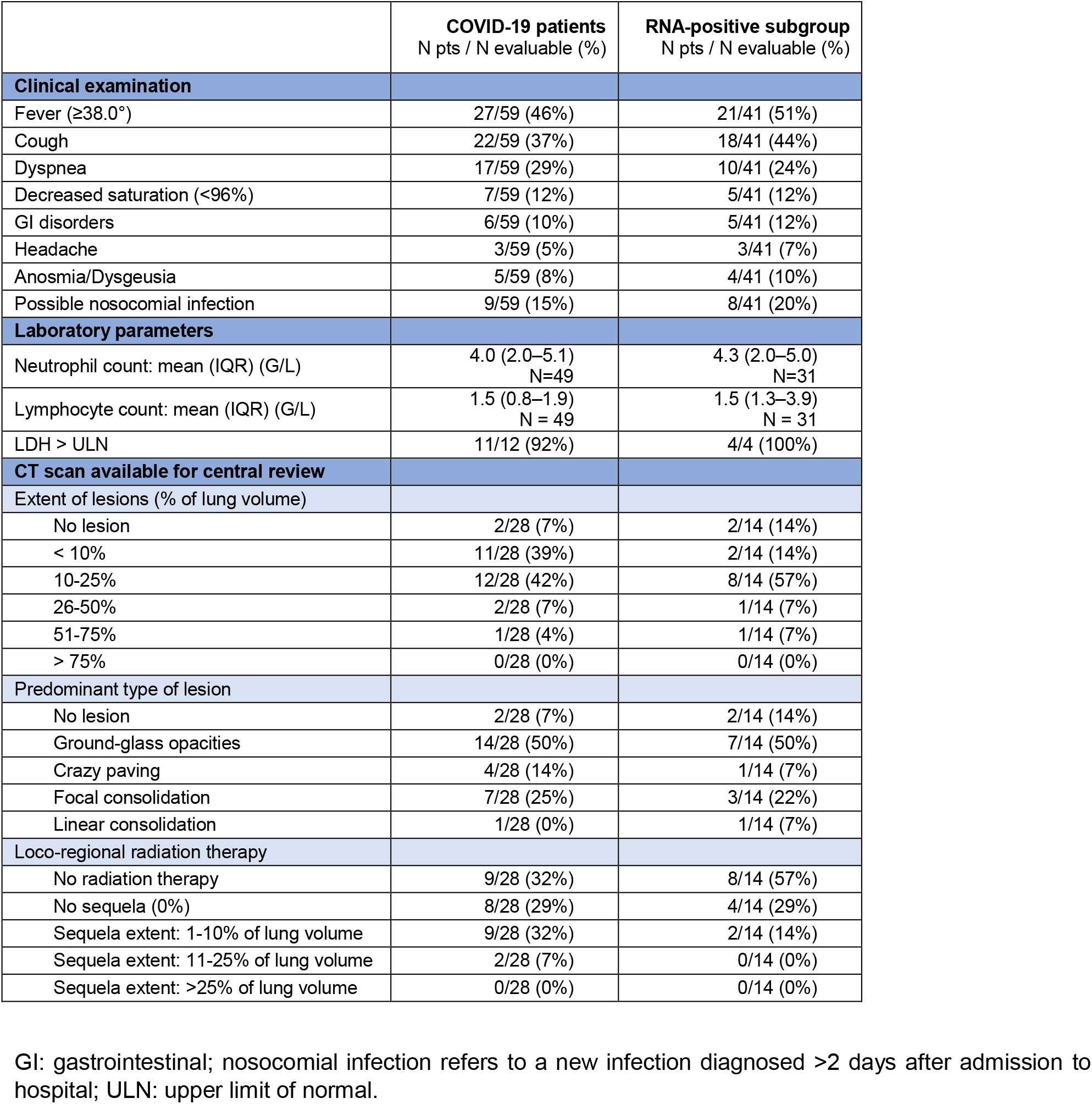
Clinical, laboratory and radiologic features at first examination.

### Outcome and prognostic factors

All patient outcomes were updated 2 days prior to this analysis. Of the 59 breast cancer patients diagnosed with COVID-19, 28 (47%) were hospitalized, while 31 (53%) returned home. Twenty-three (82%) of the 28 hospitalized patients received antibiotics and 3 (11%) received corticosteroids. No patients received hydroxychloroquine, antiviral or immunomodulating drugs as frontline treatment at admission. The use of these putative treatments, which were available whenever necessary throughout the patient’s stay in hospital and was not always available for patients hospitalized outside ICH.

None of the 17 symptom-only patients had to be hospitalized. The flow of COVID-19 patients during the course disease is shown in **Figure 2**. Four patients were transferred to ICU at diagnosis or during hospitalization. As of April 24^th^, 45 (76%) of the 59 COVID-19 patients were considered to be either recovering or cured. The outcome of 10 (17%) patients remains undetermined (most recent cases with limited follow-up), while 4 (6.7%) patients died: 2 patients were receiving later lines of treatment for metastatic breast cancer (these patients were not transferred to ICU), 1 patient had recently started first-line endocrine therapy combined with palbociclib and 1 patient was receiving neoadjuvant chemotherapy. Noteworthy, this last patient was treated with an anti-CD80/86 antibody (regulating CTLA-4 signaling). Further details on the history of the four deceased patients are available in **Table 4**.

**Figure 2:**
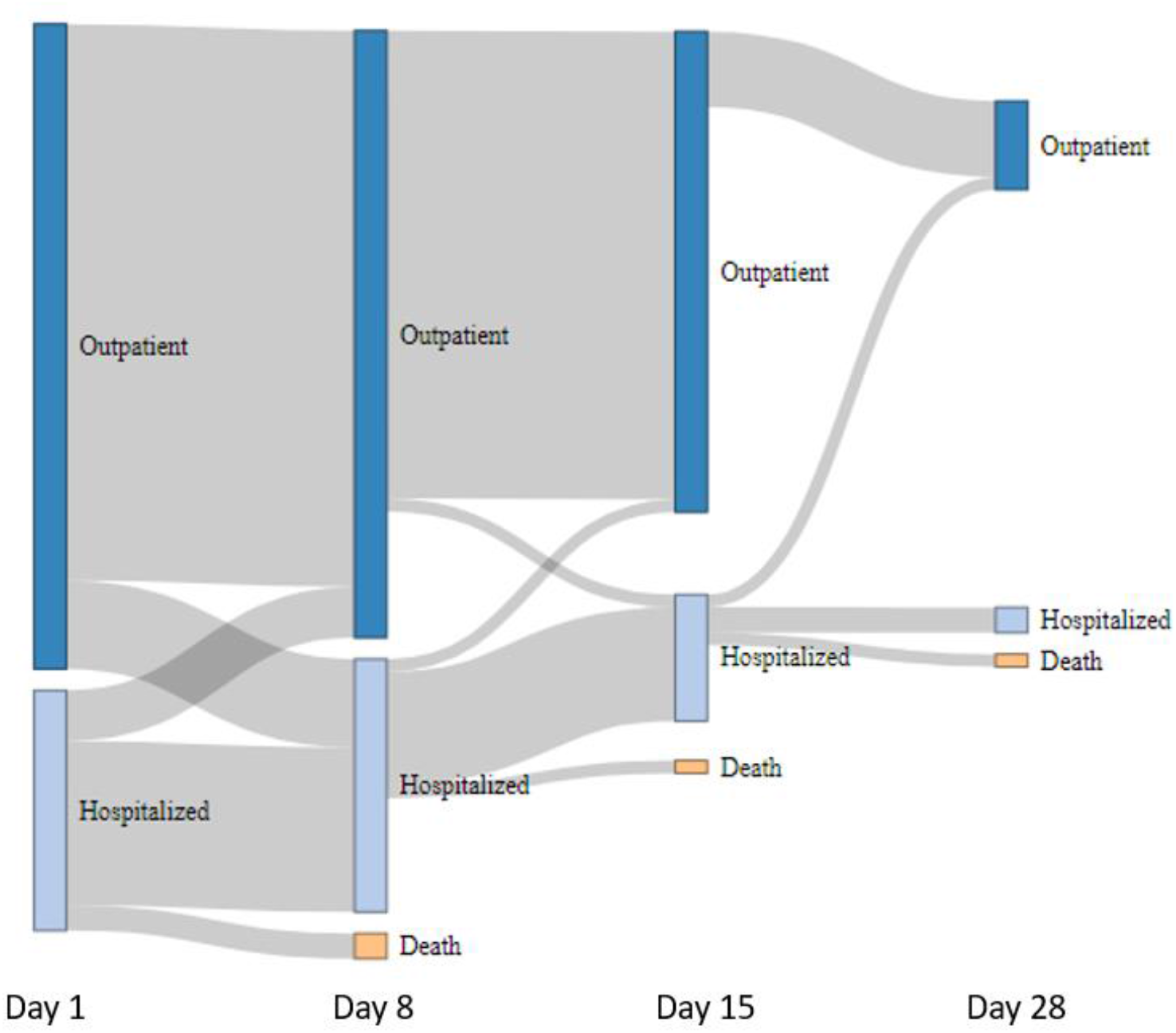
COVID-19 patients’ trajectory. Follow up consisted in clinical evaluation by phone calls scheduled at days 8, 14 and 28.

**Table 4:**
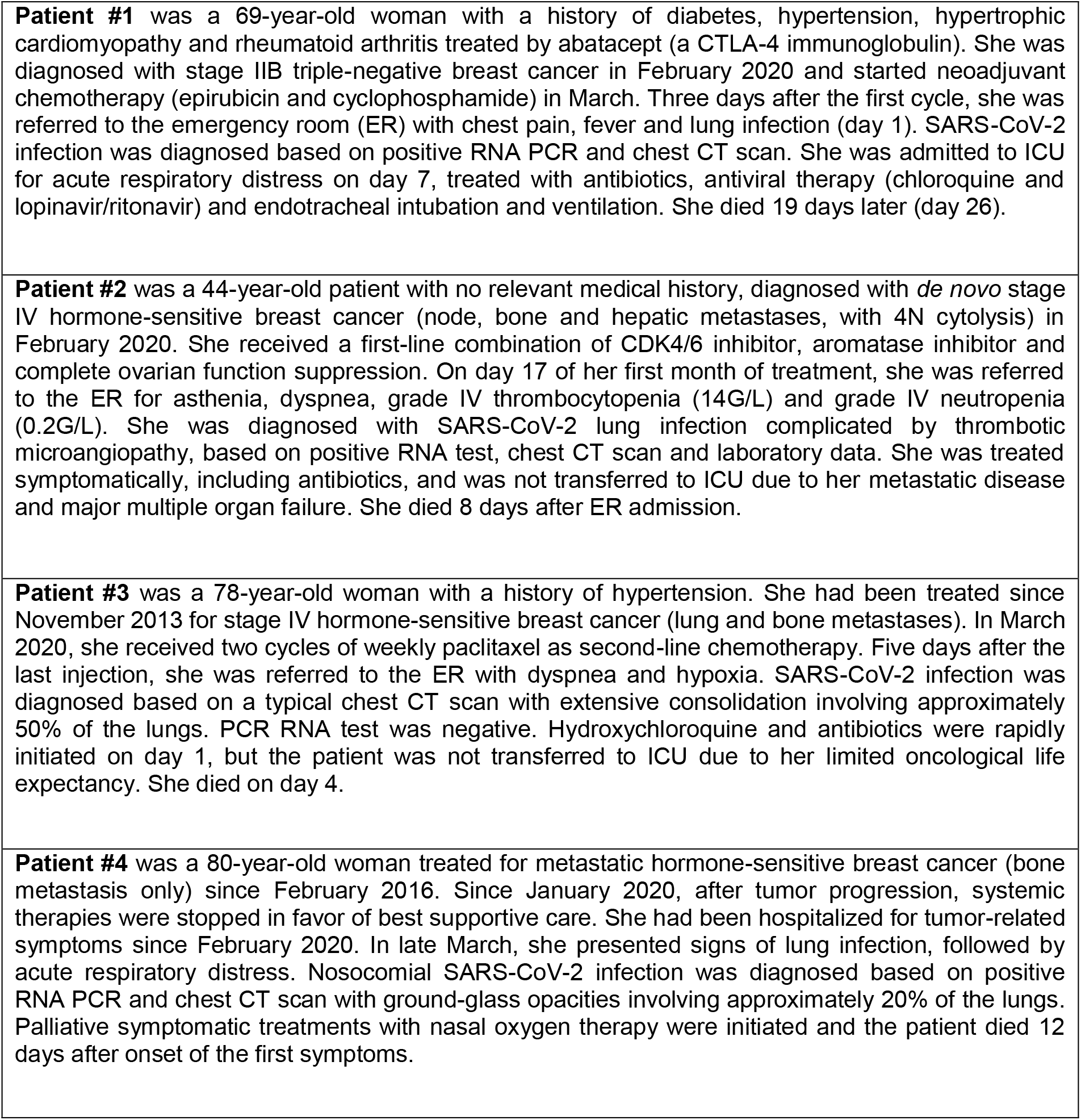
Description of COVID-19-related deaths.

An exploratory analysis of factors associated with either ICU admission and/or death in the COVID-19 population showed that, among all factors listed in Tables 1-3, only age > 70 years and hypertension were significantly associated with COVID-19 severity (both p<0.05). More specifically, the ongoing systemic treatment type (**Supplementary Table 1**), lymphopenia (<0.5G/L), neutropenia (<1G/L) and use of Angiotensin converting enzyme inhibitors or angiotensin receptor blockers had no significant prognostic impact (all p value >0.6). Age and hypertension remained as prognostic factors in the subgroup of RNA test-positive patients except that hypertension was of borderline significance. Same statistical conclusions were obtained with the analyses of time to death or ICU admission.

## Discussion

The SARS-CoV-2 outbreak is the first viral pandemic affecting cancer patients and oncology teams. To the best of our knowledge, this is the first report on COVID-19 diagnosis, signs and outcome in breast cancer patients.

While 15,600 patients were actively treated for breast cancer at Institut Curie Hospitals over the 4 months prior to the pandemic, only 59 were diagnosed with COVID-19 by either RNA test or CT scan. A recent study estimated that more than 10% of inhabitants of the greater Paris area have been infected by the SARS-CoV-2 virus [19]. While our study cannot determine the incidence of COVID-19 infection among breast cancer patients, the small number of diagnosed cases suggests that breast cancer patients do not appear to be at higher risk than the general population. This apparent low incidence could possibly be attributed to much stricter application of social distancing procedures by cancer patients, who had been informed that they may be at higher risk of severe COVID-19 infection. Prophylactic changes implemented in breast cancer care (e.g., postponement of all non-mandatory visits to ICH, changes in medical treatments, etc.) may also have contributed to further reduce the risk of SARS-CoV-2 infection. A limitation of our study is that some patients may have been treated by their family physicians or referred to local hospitals, without any notification to ICH. Although no data was available to compare COVID-19 patients to the other breast cancer patients seen at ICH, rates of high BMI and hypertension in our COVID-19 patient cohort were very similar to those reported in a recent prospective large-scale report on French breast cancer patients [20], suggesting that these comorbidities do not increase the risk of COVID-19. Our analyses showed that breast cancer patients have similar clinical and radiologic features of COVID-19 to those previously described in other reports on non-cancer COVID-19 patients. Importantly, we found no trend in favor of a relationship between a history of breast and lymph node radiation therapy, radiation therapy sequela and radiologic extent of disease or outcome. Thrombotic, cardiovascular, microvascular and dermatological events were not recorded, as their association with COVID-19 was not fully recognized when the registry was set up.

In terms of COVID-19 outcome, we observed a non-negligible mortality rate of 6.7% (4/59) among breast cancer patients diagnosed with COVID-19, with a higher mortality rate of 9.7% (4/41) in the RNA-positive subgroup. As of April 26^th^, the reported mortality rate among RNA-positive patients in the general population ranges from 18.2% in France to 5.6% in the USA and 3.7% in Germany [4]. However, these percentages reflect more testing policy more than true differences in mortality rates. As in the general population, the true infection and mortality rates could subsequently be determined by serology tests detecting an immune response to SARS-CoV-2. Nevertheless, on univariate analysis, age and hypertension were associated with disease severity rather than the extent of disease or ongoing cancer therapy. More specifically, we found no statistical relationship between ongoing chemotherapy and outcome. Overall, our data suggest that breast cancer patients share the same risk factors for severe COVID-19 as the general population. Strikingly, the only early breast cancer patient who died was concomitantly treated for a systemic disease by a CTLA-4 signalling modulator, suggesting that breast cancer *per se* is not a major contributor to COVID-19 mortality. Limitations of this analysis include the limited number of patients, a potential under-declaration due to the difficulty in identifying COVID-19 cases in outpatients who may have been referred to other hospitals. A longer follow-up of this registry may help defining more precisely the outcome of breast cancer patients with COVID-19.

## Conclusions

While lockdown lifting procedures are being discussed in most Western countries, this first report on breast cancer patients suggests that comorbidities (apart from breast cancer) should be the primary focus of attention to define patients at high risk. Further studies devoted to breast cancer patients will help to define breast cancer care for the following months, until preventive treatments, such as a vaccine, have been found.

## Data Availability

The datasets generated and analyzed during this study are not publicly available due to French HIPAA (birthdate, admission date, discharge date, date of death), but are available from the corresponding author on reasonable request.

## Declarations

### Ethics approval

The COVID-19 registry was approved by the Institut Curie institutional review board, which waived documentation of informed consent due to the observational nature of the registry.

### Consent for publication

Not applicable.

### Authors’ contributions

MF, TR, CB, SD, AN, AB, VS, YK, LC and JYP contributed to data collection and interpretation. LB and PC set up the registry and contributed to data interpretation. LC contributed to manuscript writing. PV and FCB collected the data, contributed to the analysis and wrote the manuscript. XP set up the registry, performed statistical analyses and contributed to writing of the manuscript. All authors have read and approved the final manuscript.

## Acknowledgments

*Institut Curie Breast Cancer and COVID Group:* Aurélia Alimi, Muriel Belotti, Okba Bensaoula, Ophélie Bertrand, Geoffroy Bilger, Etienne Brain, Hervé Brisse, Bruno Buecher, Laetitia Chanas, Caroline Chapus, Isabelle Charles-Massar, Pascal Chérel, Gilles Créhange, Christelle Colas, Hélène Delhomelle, Thomas Frederic-Moreau, Emmanuelle Fourme, Pierre Fumoleau, Marion Gauthier-Villars, Olivier Lantz, Sophie Lassalle, Marine Le Mentec, Florence Lerebours, Delphine Loirat, Matthieu Minsat, Pauline Moreau, Antoine de Pauw, Maël Priour, Fabien Reyal, Roman Rouzier, Mary Saad, Claire Saule, Clara Sebbag, Dominique Stoppa-Lyonnet, Anne Tardivon, Silvia Takanen, Dominique Vanjak, Marie-Charlotte Villy, Anne Vincent-Salomon, Mathilde Warcoin.

## Supplementary Methods 1

Data collected in the registry

Supplementary Methods 1: Data collected in the registry
|  |  |
| --- | --- |
| **Patient history and cancer History** | Date of birth<br>Primary Cancer<br>Treatment setting<br>Hormone receptor status (ER, PR)<br>HER2 status*<br>Previous adjuvant radiotherapy (fields)<br>Date of primary tumor diagnosis<br>Date of first metastasis<br>Number of prior lines of treatment (if stage IV)<br>Metastatic sites<br>BMI<br>Comorbidities<br>Chronic treatments<br>Steroids, NSAIDS, ACE inhibitor/ARB, Anticoagulants, Immunosuppressive drugs |
| **Symptoms and COVID-19 data** | Primary reason for registry<br>Site of COVID-19 diagnosis<br>Symptoms type<br>Date of first symptoms<br>At first diagnosis (and, if available > 30 days before):<br>Hemoglobin (g/dL)<br>Platelets counts (G/L)<br>Neutrophils counts (G/L)<br>Lymphocytes count G/L)<br>Lactate dehydrogenase (ULN)<br>CRP<br>PCT<br>Ferritin<br>Fibrinogen (g/L)<br>Prothrombin time (expressed in %)<br>SGOT (ULN)<br>SPGT (ULN)<br>Date of Chest CT-scan<br>Date of RT PCR test and results<br>SARS-CoV-2 treatment, <u>if initiated on the same day</u><br>Antibiotics<br>Corticosteroids<br>Choloroquin<br>Hydroxychloroquin<br>Other anti-viral<br>Other |
| **Radiological data** | Date of Chest CT-scan<br>Chest CT-scan results<br>Central reviewed performed<br>Extent of lesions<br>Predominant type of lesions<br>Residual toxicity of Mammary radiotherapy |
| **Healthcare** | Related to oncological care<br>Treatment received in the last 30 days<br>Date of the last treatment<br>Related to COVID-19 infection<br>Hospitalization status<br>Site of Hospitalization<br>Date of hospitalization<br>Date of discharge<br>Type of follow up<br>Oncological treatment delay |
| **Follow up calls** | Date of follow up<br>Time since the first screening<br>Follow up modality<br>Hospitalization (ICU?)<br>Outpatients<br>Other<br>Symptoms update<br>Reason to stop follow up<br>Death?<br>Date of death or last follow up |

Estrogen Receptor; PR: Progesterone Receptor; BMI: body mass index; ULN: Upper Limit of Normal range; NSAID: non-steroidal anti-inflammatory drugs; ACE: angiotensin-converting enzyme; ARB: angiotensin II receptor blockers.

## Supplementary Methods 2

### Patient flow

Starting on March 9, 2020, ICH provided information about COVID-19 symptoms and guidelines to patients by means of regular e-mail and text messages. Follow-up consultations were postponed. Consultations not requiring physical examination or detailed explanations were replaced by teleconsultations. Patients were systematically subjected to COVID-19 symptom screening and body temperature checks at the hospital entrances. Those with COVID-19 symptoms were directed to specific emergency room areas and were tested. When necessary, patients with suspected or proven COVID-19 were hospitalized in specific wards or transferred to internal or external ICU. Of note, Paris and its area did not run out of standard inpatients or ICU beds.

### Surgery

The French hospital emergency response plan (*“plan blanc*”) required postponement of all non-urgent surgery. ICH surgeons complied with the national guidelines for breast cancer patients issued by learned societies [1], included postponement of surgery for patients at high risk of severe COVID-19, in whom an alternate treatment option was available (eg, neoadjuvant endocrine therapy was proposed in elderly ER+ breast cancer patients). Patients were systematically tested for SARS-CoV-2 RNA (nasopharyngeal swabs) in the two days prior to general anesthesia, even in the absence of symptoms.

### Medical oncology

Internal guidelines were applied at ICH starting from March 9, in line with national guidelines for breast cancer patients [1]: ongoing chemotherapy was not discontinued. Neoadjuvant chemotherapy was avoided in patients with smaller tumors (< 3 cm), even including triple-negative and HER2-positive subtypes. Adjuvant chemotherapy indications were mostly maintained, but situations associated with a marginal survival benefit had to be discussed with the patients. Dose-dense anthracycline regimens were discouraged. Granulocyte colony-stimulating factor prophylaxis was systematically used for anthracycline-based regimens and docetaxel, with the docetaxel dose reduced to 75 mg/m^2^ . In patients experiencing COVID-19 symptoms (proven or unproven), the next chemotherapy cycle was delayed until 14 days after the first day of symptoms, pending the clearance of hyperthermia and dyspnea. Endocrine therapy indications and doses were not modified. Targeted therapy (including CDK4/6 and PARP inhibitors) indications and doses were not modified, except for alpelisib and everolimus. Initiation of a new line of therapy with alpelisib was discouraged, but ongoing treatment was maintained. Everolimus was discontinued in all patients, except for a handful of patients with prolonged objective response. Atezolizumab indications and doses were not modified, but close monitoring of these patients was recommended. When used as comedication for chemotherapy, corticosteroid doses were systematically reduced by one-half and were discontinued whenever possible.

### Radiation therapy

Indications and ongoing treatments were not modified, but adjuvant radiotherapy was hypofractionated whenever possible [1, 2].

### Palliative care

For patients with advanced metastatic breast cancer and/or severe pre-existing comorbidities, a dedicated COVID-19 videoconference multidisciplinary meeting was held daily to discuss the medical and ethical relevance of antiviral treatments and transfer to ICU. Palliative care for breast cancer patients during the COVID-19 pandemic was conducted according to the guidelines issued by the French society of palliative care [3].

---

**Supplementary Table 1.** Impact of systemic treatment modalities on COVID-19 severity (ICU admission or death), univariate analyses.

| Systemic treatment received within 1 month before COVID-19 diagnosis | Odds ratio | 95%CI | P value |
| --- | --- | --- | --- |
| Chemotherapy | 1.6 | [0.3-7.7] | 0.6 |
| Endocrine therapy | 0.8 | [0.1-4.7] | 0.8 |
| Targeted therapy | 0.8 | [0.1-4.3] | 0.8 |

**Supplementary Figure 1:**
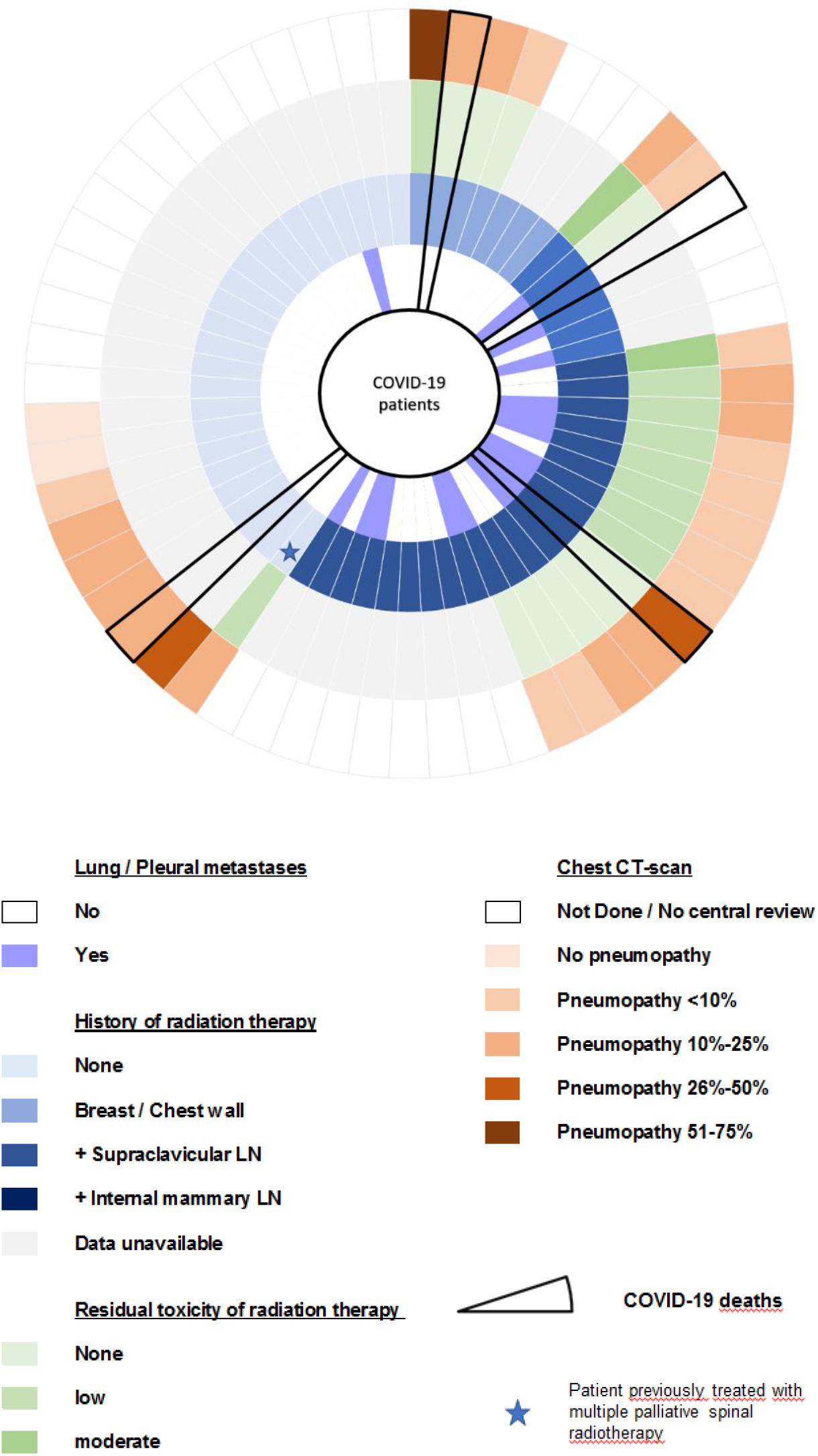
Radiation therapy and COVID-19 features Presence of pleural or lung metastases (inner circle), radiation therapy sequelae (semiquantitative estimates, green circle) and extent of COVID-19 lung disease (semiquantitative estimates, red circle) are displayed by irradiation fields (blue circle) for each of the 59 COVID-19 patients. Patients who died are surrounded in black. CT scans not available for central review are not displayed on the graph (marked as ‘not done’).

